# Vital signs as a source of racial bias

**DOI:** 10.1101/2022.02.03.22270291

**Authors:** Bojana Velichkovska, Hristijan Gjoreski, Daniel Denkovski, Marija Kalendar, Behrooz Mamandipoor, Leo Anthony Celi, Venet Osmani

**Affiliations:** Ss. Cyril and Methodius University, Faculty of Electrical Engineering and Information Technologies, Skopje, R. N. Macedonia; Harvard-MIT Health Sciences and Technology, Massachusetts Institute of Technology, Cambridge, MA, 02139, USA; Fondazione Bruno Kessler Research Institute, Trento, 38123, Italy

**Keywords:** ICU data, critical care, racial bias, ethnic bias, eICU-CRD, MIMIC-III, vital signs, ethno-race prediction

## Abstract

**Background:** racial bias has been shown to be present in clinical data, affecting patients unfairly based on their race, ethnicity and socio-economic status. This problem has the potential to be significantly exacerbated in the light of Artificial Intelligence-aided clinical decision making. We sought to investigate whether bias can be introduced from sources that are considered neutral with respect to ethnicity and race and consequently routinely used in modelling, specifically vital signs.

**Methods:** to perform our analysis, we extracted vital signs from 49,610 admissions from a cohort of adult patients during the first 24 hours after the admission to the Intensive Care Units (ICU), derived from multi-centre eICU-CRD database and single-centre MIMIC-III database, spanning over 208 hospitals and 335 ICUs. Using heart rate, SaO2, respiratory rate, systolic, diastolic, and mean blood pressure, we develop machine learning models based on Logistic Regression and eXtreme Gradient Boosting and investigate their performance in predicting patients’ self-reported race. To balance the dataset between the three ethno-races considered in our study, we use a matching cohort based on age, gender, and admission diagnosis.

**Findings:** standard machine learning models, derived solely on six vital signs can be used to predict patients’ self-reported race with AUC of 75%. Our findings hold under diverse patient populations, derived from multiple hospitals and intensive care units. We also show that oxygen saturation is a highly predictive variable, even when measured through methods other than pulse oximetry, namely arterial blood gas analysis, suggesting that addressing bias in routinely collected clinical variables will be challenging.

**Interpretation:** our finding that machine learning models can predict self-reported race using solely vital signs creates a significant risk in clinical decision making, further exacerbating racial inequalities, with highly challenging mitigation measures.

**Funding:** The funders had no role in the design of this study.

## 1 Introduction

Machine learning (ML) algorithms are being increasingly used to tackle particularly complex clinical challenges [1],[2],[3]. Looking ahead, clinical practice will benefit from game-changing approaches that can assist healthcare processes via (semi-) autonomous decision making and/or recommendations of care actions. Some of these algorithms may possibly focus on decisions where patients’ lives are at risk. Therefore, the ML algorithms that become part of clinical practice must be robust, reliable, and unbiased.

Bias can be introduced from the data or stem from the approach used in the model development pipeline, and in both cases, it can result in decision making that can be discriminatory and harmful to minority groups. Importantly, the bias stemming from data is especially critical since it propagates and even amplifies inequalities in under-represented groups.

The unequal treatment of patients based on their race has been reported in detailed studies, accompanied by indisputable evidence of bias in healthcare providers’ attitudes, expectations, and behaviour [4],[5],[6]. The problem with the presence of bias in medical practice is further amplified if considered that the bias might be taught to students, as is heavily implied in an opinion piece in [7] defining the process as a “silent curriculum”, stating “among two patients in pain waiting in an emergency department examination room, the white one is more likely to get medications and the black one is more likely to be discharged with a note documenting “narcotic-seeking behavior”. A recent study [8] illustrates racial bias in the descriptions of patients’ electronic health records (EHR), showing that black patients are 2.5 times more likely to have one or more negative descriptors in their EHR compared with white patients.

Biased medical decisions may also result from clinical trials that produce biased datasets, either obtained entirely from a single ethno-racial group or with a dominant representation of one ethno-race over others [9]. The study in [10] shows that, even though ethno-race influences response to cancer treatments and outcomes, no ethno-racial statuses are recorded in the majority of patients, and in cases of recorded ethno-race the highest represented ethno-race in melanoma, breast and lung cancer trials are white people (25.94%), followed by Asians (4.97%), and African Americans (1.08%); resulting in biased datasets with underrepresentation of particular ethnicities [11].

Working with biased datasets negatively influences the development of ML assisted applications. There have been reports of detected ethno-racial bias in medical ML applications. The study in [12] shows patients being assigned a risk score depending on their skin colour. In particular, Black patients which were placed in the same risk category as a subset of White patients, health-wise, had considerably worse symptoms. To add to the severity of the problem, the ML algorithm reduced the number of Black patients who should have been referred for complex care by more than half. Another example is an algorithm for the diagnosis of diabetic retinopathy showing poor performance in populations living outside of the location where it was developed [13].

Analysis of ethno-racial bias in ML applications can also be performed by observing the models’ performances across race-ethnicities [14]. In [15] the authors present their investigation into the performance of three severity scoring systems in four ethnicities, focusing on hospital mortality. The authors conclude that severity scores have a statistical bias since the overestimated mortalities are most notable with Hispanic and Black patients.

Considering these issues, we sought to investigate an unlikely contributor of bias in artificial intelligence (AI) algorithms, namely the vital signs. The development of ML models typically includes the data pre-processing phase where variables that could potentially introduce bias in the models, such as race-ethnicity are excluded. The objective is to prevent the algorithm from using attributes that should not be used for classification, prediction or optimisation. For example, an algorithm that sifts through curriculum vitae should not be using the gender or the race-ethnicity as input so these are routinely excluded as input when the algorithms are trained. Vital signs, such as blood pressure, heart rate, and oxygen saturation, are objective measures and considered neutral with respect to demographics, ethnicity and race and as such considered safe, unbiased candidate features, for modelling. We investigate whether seemingly bias-free data can contain information about sensitive attributes such as race-ethnicity that can be learned during training, specifically focusing on vital signs.

To perform our analysis, we rely on the eICU Collaborative Research Database [16] where we extract the vital signs from 31,849 admissions of adult patients and the MIMIC-III [17] dataset where we extract the vital signs of a cohort of 17,761 admissions of adult patients, considered in the first 24 hours after the admission. Since both cohorts are made up of predominantly Caucasian patients, we used a matching cohort based on age, gender, and admission diagnosis to balance the dataset between the three ethno-races considered in our study, namely Caucasian, African American and Hispanic. Classical ML algorithms (Logistic Regression and XGBoost) are then used to investigate whether the patients’ ethno-race can be predicted based solely on their vital signs. We also perform variable saliency analysis, to identify the extent to which specific vital signs contribute most towards the prediction of the ethno-race.

The rest of the paper is organised as follows. Section II describes the dataset, the data preparation process and the methodology used. Section III presents our results, whereas in section IV we discuss them. Section V concludes this paper and discusses our potential future work in this area.

## 2 Methods

### 2.1 Clinical data sources and study population

The eICU Collaborative Research Database (eICU-CRD), used in our study, contains data associated with 200,859 admissions collected from 335 ICUs across 208 hospitals in the US admitted between 2014 and 2015 [16]. We additionally used the MIMIC-III [17] database, which comprises data of over 40,000 patients admitted in critical care units between 2001 and 2012. From both datasets, we selected all adult patients (age 18 and over) that were alive within the first 24 hours after ICU admission that had at least one clinically valid measurement (see Appendix 2, Table I) for all the six vital signs considered for this study: heart rate, SaO2, respiratory rate, systolic, diastolic and mean blood pressure. We additionally extracted several statistical features including mean, minimum, maximum and variance. Patients that were missing admission diagnosis, age, or gender were excluded from the study.

For the eICU dataset, we used the three most dominant ethno-races: Caucasian, African American, and Hispanic. This resulted in a total of 31,849 patients, of which 27,335 were (85%) Caucasian, 3351 (11%) were African American, and 1163 were (4%) Hispanic patients. The data for the other two, Asian and Native American, was significantly lower, with 621 and 247 patients respectively, so these ethno-races weren’t used in this research. For the MIMIC-III dataset, we repeated the same selection, which resulted in 17,761 patients total, of which 15,899 (89.5%) were Caucasian, 1365 (7.7%) were African American, and 497 (2.8%) were Hispanic patients.

### 2.2 Statistical Analysis

The baseline characteristics of the patients were analysed using medians (IQRs) for continuous variables and frequencies (percentages) for categorical variables. We used the Kruskal–Wallis test (one-way ANOVA) for continuous variables and the chi-square test for categorical variables to compare ethno-racial subgroups.

### 2.3 Model development and validation

For each of the datasets, we analysed binary comparative tests, i.e., African Americans and Caucasians, Hispanics and Caucasians, Hispanics and African Americans. Additionally, to address the issue of data imbalance between ethno-races we created three matched cohorts by devising a matching process with the minority ethno-race based on three primary features: admission diagnosis (first cohort), gender (second cohort), and age (third match) (see Appendix 1). The matching process resulted in a total of 9 sets of data, for each dataset. We evaluate the performance using the Logistic Regression (LR) - a model which uses a logistic function in order to model a binary output variable given input variables. The classification is performed based on a decision threshold, which is why LR is useful when the outcome variable is binary, but the input variables are continuous; and XGBoost - uses gradient boosted trees, and is an ensemble of models that learn by correcting the errors made by existing models until no further improvements can be made. XGBoost can be prone to overfitting because when the weaker models are trained, the resulting model has high complexity, and therefore, we trained two versions of the XGBoost algorithm: a default version (with the default parameters), and an optimised version (with the parameters selected with random search).

The algorithms were internally evaluated using stratified 5-fold cross-validation, meaning the data was divided into 5 folds in a way each fold maintains the original distribution class-wise. The training of the model was performed on 4 folds, whereas the remaining fold was used to validate the model’s performance. This process was repeated 5 times, for each of the folds, and the final results were averaged over all folds.

The performance of our models was assessed by computing the area under the receiver operator characteristic curve (AUC-ROC) and the area under the precision-recall curve (AU-PRC). The AUC-ROC shows how well the model is capable of distinguishing between the classes and is plotted with the true positive rate (recall) on the y-axis and the false positive rate (FPR) on the x-axis. The recall represents the model’s ability to correctly classify positive samples as positive. The FPR shows the samples incorrectly classified by the model as positives out of all negative samples. The AU-PRC shows the trade-off between precision and recall. The precision represents the model’s ability not to classify a negative sample as positive.

## 3 Results

The results for all matched cohorts for both datasets showed similar performance for all models across all comparative tests, with the second matched cohort that provided the best results. Therefore, the results presented in this section are from the second matched cohort. The patient baseline characteristics in each of the three comparative tests for the second matched cohort are summarised in Table I. The baseline characteristics for the first and third matched cohort are provided in Appendix 1).

**Table I.** Cohort characteristics table for each comparative test in the second matched cohort (eICU-CRD and MIMIC-III datasets). Continuous variables are represented as medians with interquartile ranges.

| Variables | African American and Caucasian |  |  | Hispanic and Caucasian |  |  | Hispanic and African American |  |  |
| --- | --- | --- | --- | --- | --- | --- | --- | --- | --- |
|  | African American | Caucasian | p-value | Hispanic | Caucasian | p-value | Hispanic | African American | p-value |
| **eICU-CRD** |  |  |  |  |  |  |  |  |  |
| **Patients** | **3346** | **3346** | **-** | **1161** | **1161** | **-** | **1163** | **1163** | **-** |
| **Gender (male)** | **1801 (53.83%)** | **1801 (53.83%)** | **-** | **674 (58.05%)** | **674 (58.05%)** | **-** | **676 (58.13%)** | **676 (58.13%)** | **-** |
| **Age** | **58 [48, 67]** | **61 [52, 71]** | **< 0.001** | **62 [51, 73]** | **63 [53, 74]** | **0.022** | **62 [51, 73]** | **60 [50, 69]** | **0.011** |
| **Heart rate** | **87 [75, 100]** | **84 [73, 97]** | **0.007** | **84 [73, 97]** | **84 [74, 96]** | **0.811** | **84 [73, 97]** | **86 [75, 99]** | **0.019** |
| **Oxygen Saturation** | **99 [97, 100]** | **98 [96, 99]** | **<0.001** | **98 [96, 100]** | **98 [96, 99]** | **0.005** | **98 [96, 100]** | **99 [97, 100]** | **0.002** |
| **Respiration rate** | **18 [15, 23]** | **19 [15, 23]** | **0.729** | **19 [15, 23]** | **19 [15, 23]** | **0.438** | **19 [15, 23]** | **19 [15, 23]** | **0.610** |
| **Systolic blood pressure** | **126 [109, 146]** | **121 [106, 139]** | **<0.001** | **124 [109, 140]** | **122 [106, 139]** | **0.018** | **124 [109, 140]** | **124 [108, 143]** | **0.852** |
| **Diastolic blood pressure** | **62 [53, 71]** | **59 [51, 68]** | **0.506** | **59 [50, 68]** | **58 [51, 67]** | **0.133** | **59 [50, 68]** | **61 [53, 71]** | **< 0.001** |
| **Mean blood pressure** | **82 [72, 93]** | **79 [70, 90]** | **0.005** | **80 [70, 90]** | **79 [70, 90]** | **0.635** | **80 [70, 90]** | **81 [72, 92]** | **0.012** |
| **MIMIC-III** |  |  |  |  |  |  |  |  |  |
| **Patients** | **1333** | **1333** | **-** | **485** | **485** | **-** | **496** | **496** | **-** |
| **Gender (male)** | **606 (45.46%)** | **606 (45.46%)** | **-** | **301 (62.06%)** | **301 (62.06%)** | **-** | **312 (62.9%)** | **312 (62.9%)** | **-** |
| **Age** | **61 [51, 72]** | **63 [52, 73]** | **0.349** | **55 [43, 68]** | **56 [46, 69]** | **0.148** | **54 [43, 67]** | **57 [46, 68]** | **0.041** |
| **Heart rate** | **87 [75, 100]** | **85 [74, 96]** | **0.001** | **89 [78, 103]** | **86 [75, 97]** | **0.016** | **89 [78, 103]** | **87 [75, 99]** | **0.973** |
| **Oxygen Saturation** | **99 [97, 100]** | **99 [96, 100]** | **0.01** | **99 [97, 100]** | **99 [97, 100]** | **0.697** | **99 [97, 100]** | **99 [97, 100]** | **0.237** |
| **Respiration** | **18 [14, 22]** | **18 [14, 21]** | **0.52** | **17 [14, 21]** | **18 [14, 22]** | **0.853** | **17 [14, 21]** | **18 [14, 22]** | **0.044** |

| rate |  |  |  |  |  |  |  |  |  |
| --- | --- | --- | --- | --- | --- | --- | --- | --- | --- |
| Systolic blood pressure | 119 [103, 138] | 114 [101, 130] | < 0.001 | 117 [104, 134] | 113 [100, 129] | 0.003 | 117 [104, 134] | 121 [105, 139] | 0.013 |
| Diastolic blood pressure | 61 [54, 70] | 57 [50, 65] | < 0.001 | 62 [54, 71] | 58 [51, 67] | 0.001 | 62 [54, 71] | 63 [54, 72] | 0.314 |
| Mean blood pressure | 80 [70, 92] | 76 [68, 86] | < 0.001 | 80 [71, 91] | 76 [67, 86] | < 0.001 | 80 [71, 91] | 81 [71, 93] | 0.982 |

### 3.1 eICU-CRD Results

Table II summarises the eICU-CRD results from the AUC-ROC and AU-PRC for all comparative tests performed with the second matched cohort and each of the three algorithms: LR, XGBoost (with default parameters), and optimised XGBoost (with optimised parameters). The comparative test between Hispanic and Caucasian patients shows the lowest results across all algorithms. On the other hand, the best results were obtained in the comparative test between African American and Caucasian patients, with the highest AUC performance at 0.75 ± 0.019. The results additionally show that XGBoost performed better than LR across all tests. The optimised XGBoost performed better than the default XGBoost, due to the simplification of the ensemble models used, which removed the model’s overfitting.

**Table II.** AUC and AP across all comparative tests for the second matched cohort and each of the three algorithms used (eICU-CRD). The values are represented with mean and standard deviation.

|  | African American and Caucasian |  | Hispanic and Caucasian |  | Hispanic and African American |  |
| --- | --- | --- | --- | --- | --- | --- |
| Model | AUC | AP | AUC | AP | AUC | AP |
| LR | $0.64 \pm 0.016$ | $0.62 \pm 0.059$ | $0.63 \pm 0.056$ | $0.58 \pm 0.042$ | $0.70 \pm 0.051$ | $0.62 \pm 0.064$ |
| XGBoost Default | $0.71 \pm 0.023$ | $0.66 \pm 0.071$ | $0.64 \pm 0.04$ | $0.60 \pm 0.062$ | $0.69 \pm 0.041$ | $0.61 \pm 0.053$ |
| XGBoost Optimised | **$0.75 \pm 0.019$** | **$0.70 \pm 0.089$** | **$0.67 \pm 0.047$** | **$0.63 \pm 0.065$** | **$0.72 \pm 0.045$** | **$0.65 \pm 0.081$** |

The AUC-ROC and AU-PRC are displayed in Table III. Between the three comparative tests, the graphs show the best results in the comparison between African Americans and Caucasians. Additionally, the AUC-ROC clearly displays the difference between the performance of the three algorithms, illustrating that the optimised XGBoost provides the best ethno-race prediction. The AUC-ROC and AU-PRC for the first and third matched cohorts are provided in Appendix 3 and 4.

**Table III.**
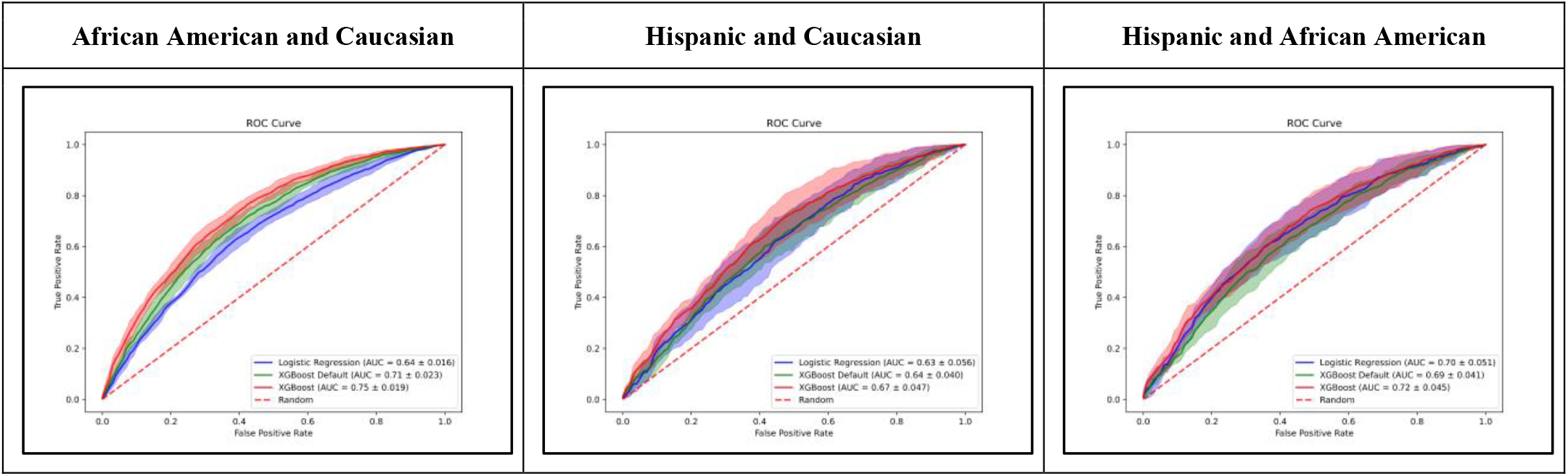
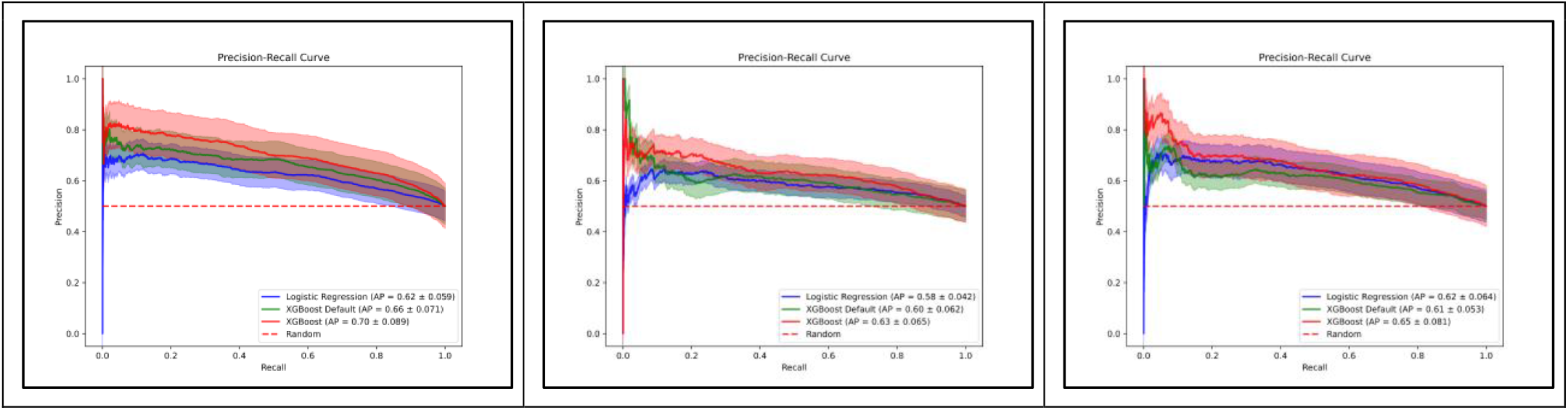
AUC-ROC and AU-PRC for the second matched cohort across all comparative tests (eICU-CRD)

### 3.2 MIMIC-III Results

Table IV summarises the MIMIC-III results from the AUC-ROC and AU-PRC for all comparative tests performed with the second matched cohort and each of the three algorithms: LR, XGBoost (with default parameters), and optimised XGBoost (with optimised parameters). The best results were obtained in the comparative test between African American and Caucasian patients, with the highest AUC performance at 0.65 ± 0.021, whereas the comparative test between Hispanic and Caucasian patients, in summary, shows the lowest results. XGBoost performed better than LR in all, except the comparison between Hispanic and Caucasian patients - the better performance of LR here can be due to potential linearity in the comparison, or the number of patients in the cohort.

The AUC-ROC and AU-PRC are displayed in Table V. Between the three comparative tests, the graphs show the best results in the comparison between African Americans and Caucasians. The other two comparative tests show marginally worse results, which was expected considering the low number of Hispanic patients remaining after the selection criteria were applied to the MIMIC-III dataset.

**Table IV.** AUC and AP across all comparative tests for the second matched cohort and each of the three algorithms used (MIMIC-III). The values are represented with mean and standard deviation.

|  | African American and Caucasian |  | Hispanic and Caucasian |  | Hispanic and African American |  |
| --- | --- | --- | --- | --- | --- | --- |
| Model | AUC | AP | AUC | AP | AUC | AP |
| LR | $0.62 \pm 0.02$ | $0.61 \pm 0.055$ | $0.62 \pm 0.033$ | $0.57 \pm 0.048$ | $0.62 \pm 0.036$ | $0.59 \pm 0.06$ |
| XGBoost Default | $0.62 \pm 0.017$ | $0.61 \pm 0.067$ | $0.53 \pm 0.035$ | $0.55 \pm 0.040$ | $0.61 \pm 0.035$ | $0.56 \pm 0.05$ |
| XGBoost Optimised | $0.65 \pm 0.021$ | $0.64 \pm 0.075$ | $0.60 \pm 0.026$ | $0.58 \pm 0.062$ | $0.64 \pm 0.038$ | $0.58 \pm 0.056$ |

**Table V.**
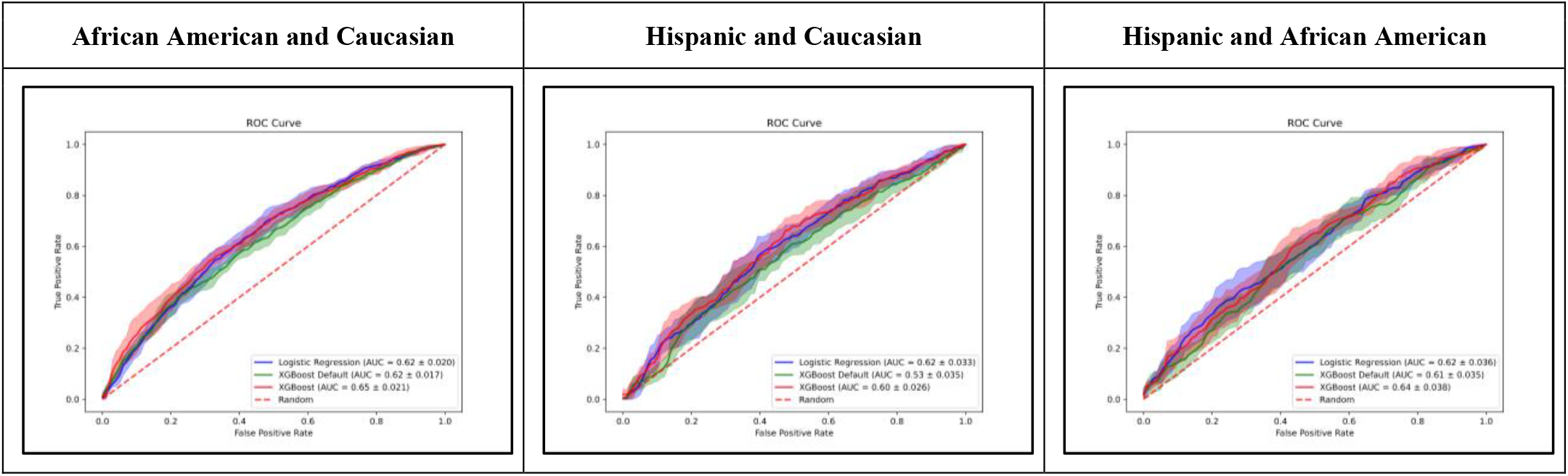
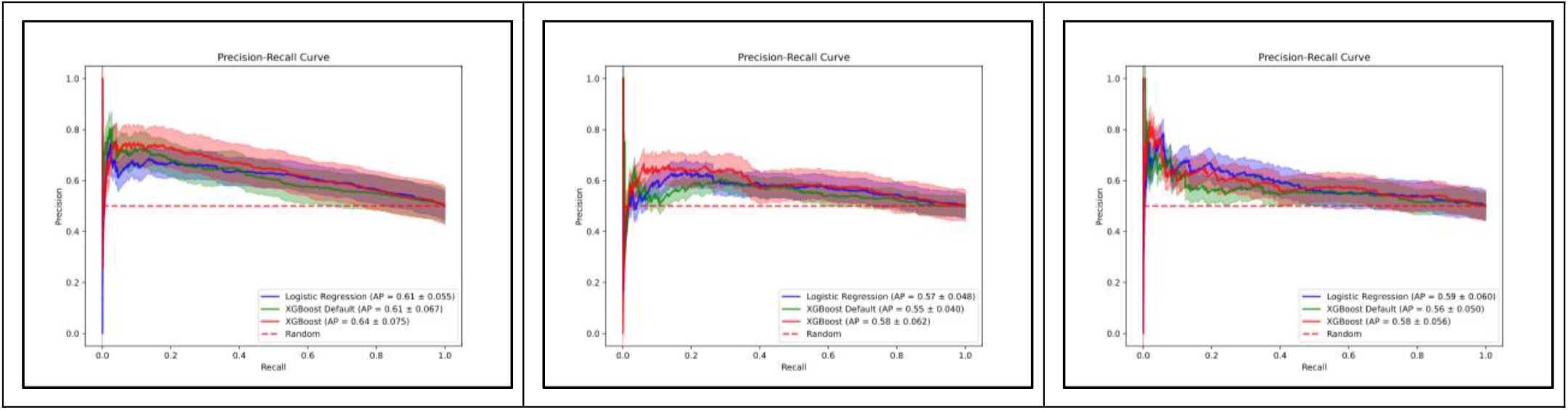
AUC-ROC and AU-PRC for the second matched cohort across all comparative tests (MIMIC-III)

### 3.3 Variable importance (eICU-CRD and MIMIC-III)

In order to understand our results better, we wanted to analyse which variables contributed most to the prediction outcome. Since the optimised XGBoost algorithms provided the best results in all cases, we inspected the variable importance of the optimised XGBoost model for each comparative test. We used the SHAP (SHapley Additive exPlanations) values to tell us how much each input variable in the model contributed to the prediction. The SHAP beeswarm plots across all comparative tests for the second matched cohort in the eICU-CRD are displayed in Table VI. When comparing African American and Caucasian patients, we see that the beeswarm plot shows the oxygen saturation carries high importance for the XGBoost classifier. This was true also for the MIMIC-III cohort

**Table VI.**
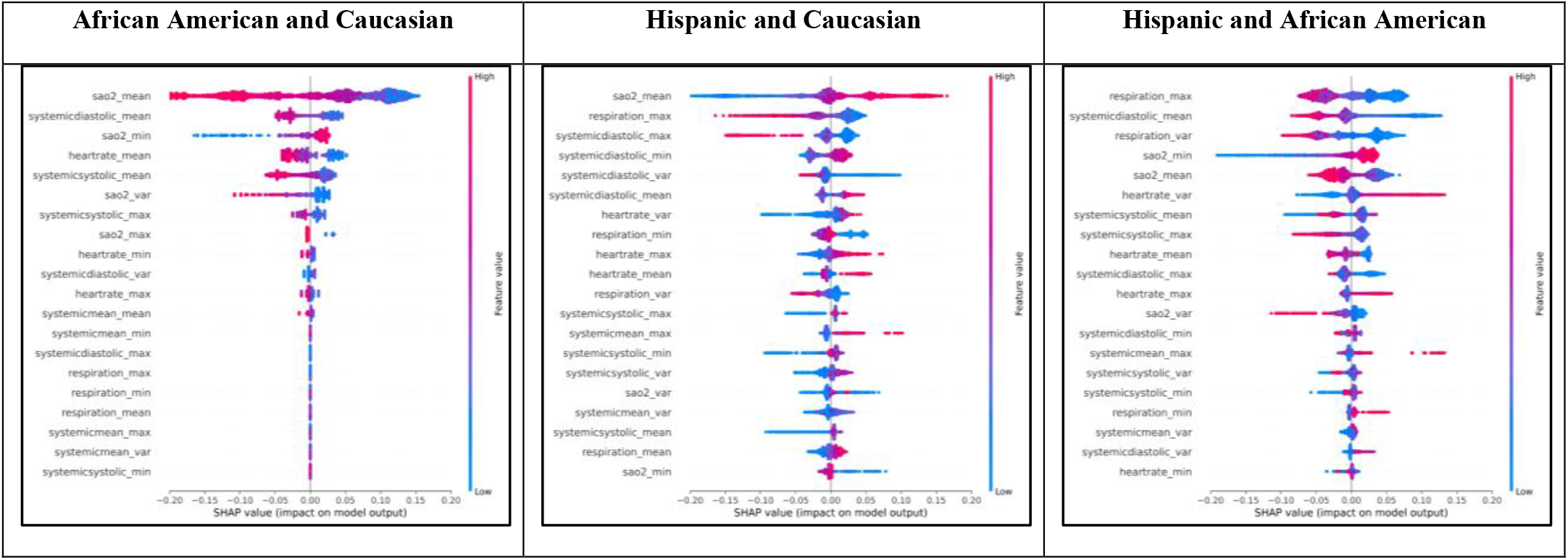
SHAP beeswarm variable importance for the second matched cohort across all comparative tests (eICU-CRD)

The mean value of the oxygen saturation is important in the comparison of Hispanics and Caucasians, as well. In this comparison, the maximum respiration is also important to the model. When comparing Hispanic and African American patients the respiration maximum and variance were among the most relevant variables for the model, which was also the case when comparing Hispanic with Caucasian patients.

We performed the variable analysis on the MIMIC-III dataset, and we again focused on the results provided by the optimised XGBoost (in spite of LR performing better in the case of Hispanic and Caucasian patients, the difference in the result is not significant). The SHAP beeswarm plots across all comparative tests for the second matched cohort are displayed in Table VII. The oxygen saturation and diastolic blood pressure are among the topmost important features in the comparative tests between the African American and Caucasian patients, and Hispanic and Caucasian patients. When comparing African American and Hispanic patients the heart rate variables significantly contribute to the decisions of the model.

**Table VII.**
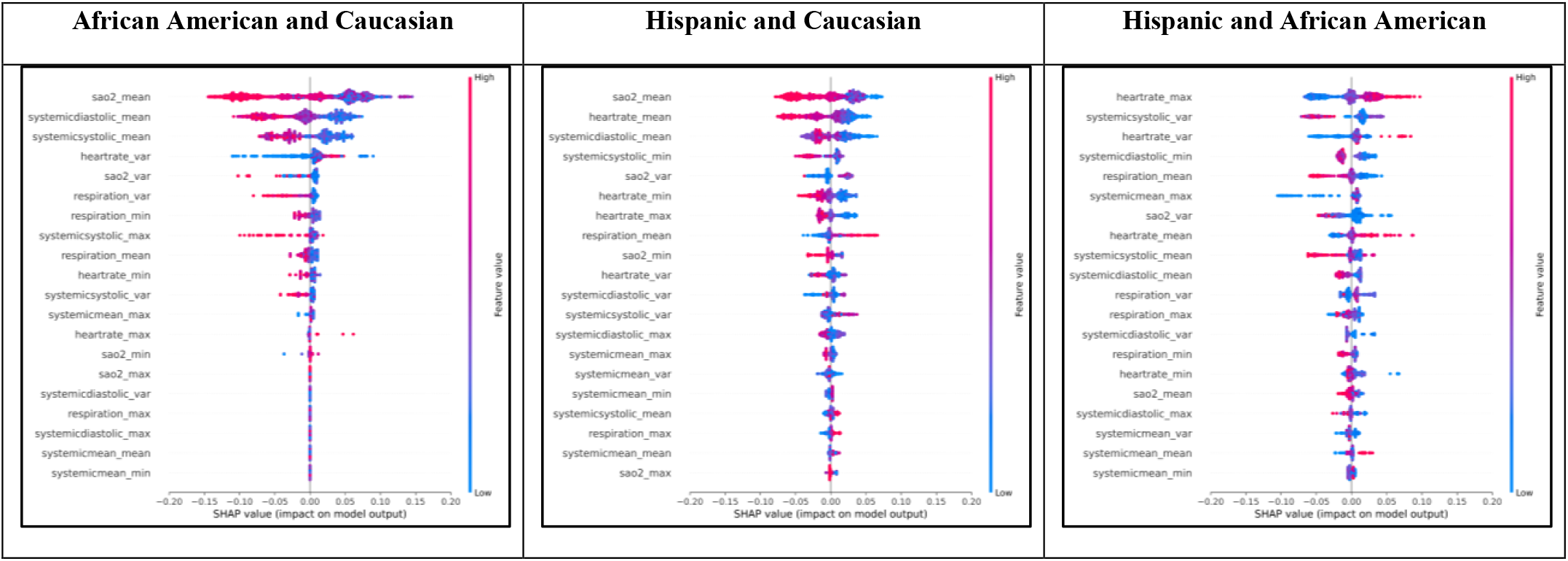
SHAP beeswarm variable importance for the second matched cohort across all comparative tests (MIMIC-III)

## 4 Discussion

From the eICU-CRD results, it can be observed that across all comparative tests the optimised XGBoost performed well with respect to identifying the race-ethnicity from the vital signs after matching for age, gender and diagnosis. When comparing African Americans and Caucasians, the AUC-ROC and AU-PRC showed the best predictive performance of the algorithms compared to the other two comparative tests. Additionally, in comparing African Americans and Caucasians the standard deviations of each algorithm are lower compared to the other two comparative tests. This outcome can be a result of the number of patients representative of each ethno-race in the comparative tests; namely, during the matching process, we can see the resulting data for the African American and Caucasian comparative test has approximately thrice the number of patients in the Hispanics and Caucasians and Hispanics and African Americans patient pools. However, the significantly larger number of patients does not provide a pronounced difference in the results, signalling that it might be that the selected variables do not offer additional insight into the distinction between two ethno-races. We observed lower model performance on the MIMIC-III dataset, in part due to the lower number of patients selected from the MIMIC-III. However, the MIMIC-III results follow the same trend, i.e., the best results were obtained in the second matched cohort (where the gender was the primary match feature), the comparative tests of African American and Caucasian patients showed best results across all matched cohorts, and the oxygen saturation is the most important feature for the African American and Caucasian patients, and the Hispanic and Caucasian patients.

While the results show that a definite division between each ethno-race cannot be obtained, the success of the model in classifying two out of three patients correctly cannot be accidental. The models are using only the information from patients’ vital signs, suggesting that routinely collected information in the first 24 hours of admission, excluding demographic information, can provide information about the race-ethnicity even when such information is removed from the dataset.

These results could be influenced by several factors. Firstly, the bias can be introduced from the socio-economic factors, that is patients from ethno-racial minorities tend to have poorer preventive care, and consequently once admitted into the ICU they can be in significantly worse condition compared to other ethno-races even with the same admission diagnosis that we control for. Additionally, there is potential for underlying issues regarding the accuracy of medical equipment across different populations. For example, it was shown that pulse oximeters were inaccurate for certain races, which came to light during the COVID crisis, resulting in hidden hypoxemia among patients of colour [18]. This was further investigated, and studies showed that oxygen saturation levels had greater variability in patients who identified as African American, followed by Hispanic, Asian, and lastly, Caucasian patients. While our saliency analysis showed oxygen saturation as an important variable, this may not fully explain our finding as we used measures from arterial oxygen saturation measured directly through blood gas analysis, rather than pulse oximetry, where the discrepancies were found.

## 5 Conclusions

ML applications in medicine show success in performing tasks such as diagnosis and prognostication at the same level as experts. However, various studies have demonstrated how ML applications can be biased and therefore affect patients unfairly, favouring one group of patients over the other depending on race, income, etc. The bias observed in ML applications can be a result of the model development or be introduced by the data used for training algorithms. Therefore, we focused on analysing the presence of ethno-racial bias in clinical data, by investigating if vital signs could give ML models enough information to determine a patient’s ethno-race correctly. We compared the performance of three separate models in three distinct matched cohorts for two different datasets. Due to the similarity of the results between the cohorts, we focused on further analysing the results obtained from one of the cohorts. Our results show that two of three patients in all comparative tests have their ethno-race correctly identified, and the most important variables in the model decisions proved to be oxygen saturation and respiration.

## Data Availability

The datasets analyzed in the current study are publicly available in the MIMIC-III repository
(https://mimic.physionet.org/) and eICU-CRD repository (https://eicu-crd.mit.edu/).

https://mimic.physionet.org/

https://eicu-crd.mit.edu/

## Acknowledgement

This research was supported by the WideHealth project - EU Horizon 2020, under grant agreement No 952279.

## Ethical approval and consent to participate

The data in MIMIC-III was previously de-identified, and the institutional review boards of the Massachusetts Institute of Technology (No. 0403000206) and Beth Israel Deaconess Medical Center (2001-P-001699/14) both approved the use of the database for research. The analysis using the eICU-CRD is exempt from institutional review board approval due to the retrospective design, lack of direct patient intervention, and the security schema, for which the re-identification risk was certified as meeting safe harbor standards by an independent privacy expert (Privacert, Cambridge, MA) (Health Insurance Portability and Accountability Act Certification no. 1031219-2). All experiments were performed in accordance with relevant guidelines and regulations.

## Availability of data and materials

The datasets analyzed in the current study are publicly available in the MIMIC-III repository (https://mimic.physionet.org/) and eICU-CRD repository (https://eicu-crd.mit.edu/).

## 1. Appendix 1 Matching cohort

A matched cohort in our case represents a dataset created of pairs of patients from two different ethno-racial groups, who may differ with respect to their individual vital signs, however, are matched through the same specific baseline characteristics, namely age, gender and admission diagnosis. From the dataset with three ethno-races present, we created three ethno-race-based divisions for matching: African Americans vs. Caucasians, Hispanics vs. Caucasians, Hispanics vs. African Americans. For each of the three baseline characteristics (age, gender, diagnosis), we created three feature-prioritising processes for matching, each taking on one of the features as a priority in the matching process. The matching process sought to match each of the patients from the minority class with a patient from the majority class, initially and if possible, on all three features, and if there were no matches made where all three features corresponded, then for the remaining unmatched patients the matching was based on the prioritised feature.

- Match 1: The first match process prioritised the diagnosis, therefore the matching between the two ethno-racial groups followed three stages: the patients were matched on three features, then if no match was made the patients were matched on the combination of diagnosis-gender equality or diagnosis-age equality, and lastly, if no matches were found in the second stage the match was made on diagnosis only.
- Match 2: The second match process prioritised the gender feature, therefore the matching between the two ethno-racial groups followed three stages: the patients were matched on three features, then if no match was made the patients were matched on the combination of gender-age equality or gender-diagnosis equality, and lastly, if no matches were found in the second stage the match was made on gender only.
- Match 3: The third match process prioritised the age, therefore the matching between the two ethno-racial groups followed three stages: the patients were matched on three features, then if no match was made the patients were matched on the combination of age-diagnosis equality or age-gender equality, and lastly, if no matches were found in the second stage the match was made on age only. In each of the three matching processes, if by the end a patient from the minority class had no matched patient from the majority class, then the patient from the minority class was dropped.

The process provided a total of nine datasets, where each of the three ethno-race-based divisions were combined with each of the three feature-prioritising processes. The details on the patients matched and the resulting number of patients per cohort in each of the nine perfectly-balanced datasets is given in Table I.

**Table I.** The number of patient pairs matched through each matching process (and corresponding feature groups) across each comparative test (eICU-CRD and MIMIC-III)

|  | Matched on | African American and Caucasian | Hispanic and Caucasian | Hispanic and African American |
| --- | --- | --- | --- | --- |
| **eICU-CRD** |  |  |  |  |
| **Match 1** | **All three features** | 2095 (62.6%) | 837 (72%) | 327 (29.3%) |
|  | **Diagnosis gender** | 1189 (35.5%) | 321 (27.6%) | 727 (65.2%) |
|  | **Diagnosis age** | 46 (1.4%) | 4 (0.4%) | 26 (2.3%) |
|  | **Diagnosis** | 14 (0.5%) | 0 (0%) | 36 (3.2%) |
|  | **Total** | 3344 | 1162 | 1116 |
| **Match 2** | **All three features** | 2095 (62.6%) | 837 (72.1%) | 327 (28.1%) |
|  | **Gender age** | 844 (25.2%) | 231 (19.9%) | 533 (45.8%) |
|  | **Gender diagnosis** | 407 (12.2%) | 93 (8%) | 303 (26.1%) |
|  | **Gender** | 0 (0%) | 0 (0%) | 0 (0%) |
|  | **Total** | 3346 | 1161 | 1163 |
| **Match 3** | **All three features** | 2095 (62.6%) | 837 (72.1%) | 327 (28.2%) |
|  | **Age diagnosis** | 22 (0.7%) | 4 (0.3%) | 26 (2.3%) |
|  | **Age gender** | 1230 (36.7%) | 320 (27.6%) | 792 (68.4%) |
|  | **Age** | 0 (0%) | 0 (0%) | 13 (1.1%) |
|  | **Total** | 3347 | 1161 | 1158 |
| **MIMIC-III** |  |  |  |  |
| **Match 1** | **All three features** | 313 (34.1%) | 115 (34.02%) | 15 (6.15%) |
|  | **Diagnosis gender** | 541 (58.93%) | 199 (58.88%) | 185 (75.82%) |
|  | **Diagnosis age** | 15 (1.63%) | 4 (1.18%) | 3 (1.23%) |
|  | **Diagnosis** | 49 (5.34%) | 20 (5.92%) | 41 (16.8%) |
|  | **Total** | 918 | 338 | 244 |
| **Match 2** | **All three features** | 313 (23.48%) | 115 (23.71%) | 15 (3.02%) |
|  | **Gender age** | 909 (68.19%) | 328 (67.63%) | 402 (81.05%) |
|  | **Gender diagnosis** | 111 (8.33%) | 42 (8.66%) | 68 (13.71%) |
|  | **Gender** | 0 (0%) | 0 (0%) | 11 (2.22%) |
|  | **Total** | 1333 | 485 | 496 |
| **Match 3** | **All three features** | 313 (23.48%) | 115 (23.71%) | 15 (3.1%) |
|  | **Age diagnosis** | 5 (0.38%) | 3 (0.62%) | 3 (0.62%) |
|  | **Age gender** | 1015 (76.14%) | 367 (75.67%) | 451 (93.18%) |
|  | **Age** | 0 (0%) | 0 (0%) | 15 (3.1%) |
|  | **Total** | 1333 | 485 | 484 |

**Table II.**
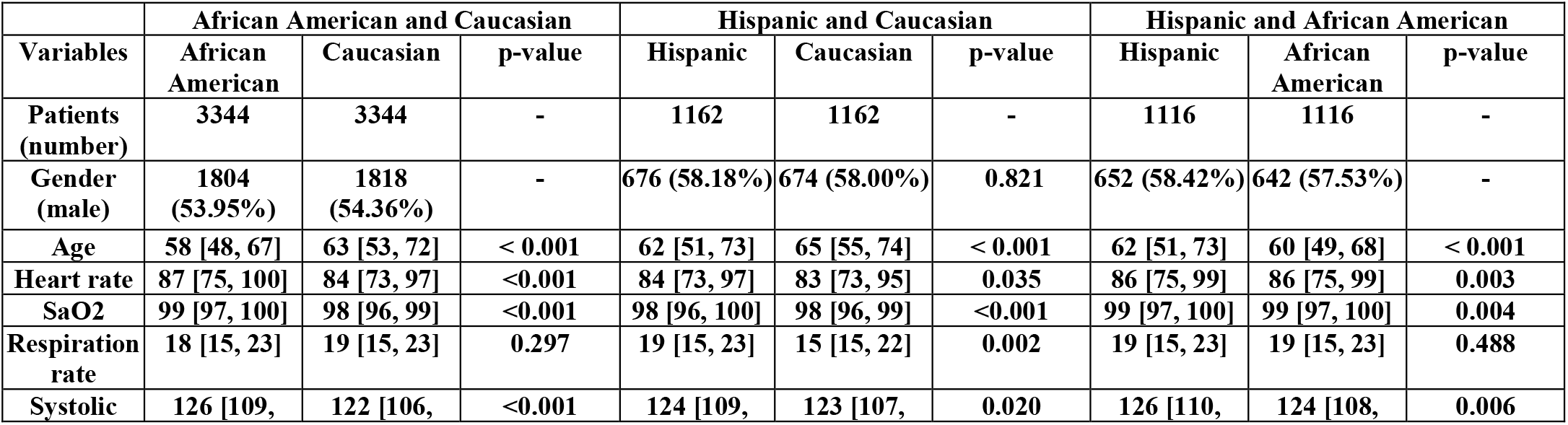
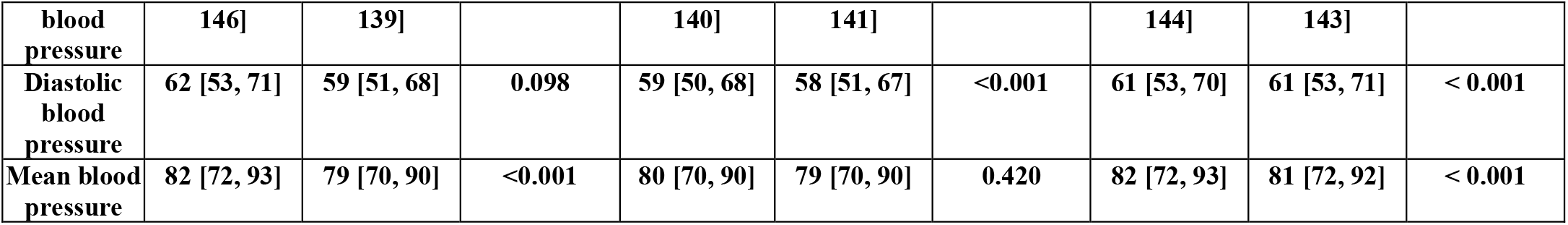
Cohort characteristics table for each comparative test in the first matched cohort (eICU-CRD). Continuous variables are represented as medians and IQRs.

**Table III.** Cohort characteristics table for each comparative test in the third matched cohort (eICU-CRD). Continuous variables are represented as medians and IQRs.

| eICU-CRD | African American and Caucasian |  |  | Hispanic and Caucasian |  |  | Hispanic and African American |  |  |
| --- | --- | --- | --- | --- | --- | --- | --- | --- | --- |
| Variables | African American | Caucasian | p-value | Hispanic | Caucasian | p-value | Hispanic | African American | p-value |
| Patients (number) | 3347 | 3347 | - | 1161 | 1161 | - | 1158 | 1158 | - |
| Gender (male) | 1802 (53.84%) | 1810 (54.08%) | - | 674 (58.05%) | 674 (58.05%) | 0.821 | 672 (58.03%) | 659 (56.91%) | - |
| Age | 58 [48, 67] | 58 [48, 67] | 0.884 | 62 [51, 73] | 62 [51, 73] | 0.801 | 62 [51, 73] | 62 [51, 73] | 0.535 |
| Heart rate | 87 [75, 100] | 85 [74, 98] | 0.008 | 84 [73, 97] | 84 [74, 96] | 0.315 | 84 [73, 97] | 86 [75, 99] | 0.084 |
| SaO2 | 99 [97, 100] | 98 [96, 99] | <0.001 | 98 [96, 100] | 98 [96, 99] | 0.001 | 98 [96, 100] | 99 [97, 100] | 0.002 |
| Respiration rate | 18 [15, 23] | 19 [15, 23] | 0.198 | 19 [15, 23] | 18 [15, 23] | 0.746 | 19 [15, 23] | 19 [15, 23] | 0.490 |
| Systolic blood pressure | 126 [109, 146] | 121 [106, 138] | <0.001 | 124 [109, 140] | 122 [106, 139] | 0.006 | 124 [109, 140] | 125 [108, 144] | 0.183 |
| Diastolic blood pressure | 62 [53, 71] | 60 [52, 69] | 0.004 | 59 [50, 68] | 59 [51, 68] | 0.253 | 59 [50, 68] | 61 [52, 70] | 0.002 |
| Mean blood pressure | 82 [72, 93] | 80 [70, 90] | < 0.001 | 80 [70, 90] | 79 [70, 90] | 0.698 | 80 [70, 90] | 81 [72, 92] | < 0.001 |

**Table IV.** Cohort characteristics table for each comparative test in the first matched cohort (MIMIC-III). Continuous variables are represented as medians and IQRs.

| MIMIC-III | African American and Caucasian |  |  | Hispanic and Caucasian |  |  | Hispanic and African American |  |  |
| --- | --- | --- | --- | --- | --- | --- | --- | --- | --- |
| Variables | African American | Caucasian | p-value | Hispanic | Caucasian | p-value | Hispanic | African American | p-value |
| Patients (number) | 918 | 918 | - | 338 | 338 | - | 244 | 244 | - |
| Gender (male) | 417 (45.42%) | 443 (48.26%) | - | 215 (63.61%) | 213 (63.02%) | - | 158 (64.75%) | 142 (58.2%) | - |
| Age | 62 [51, 73] | 66 [56, 77] | 0.014 | 55 [44, 68] | 63 [51, 74] | < 0.001 | 54 [42, 69] | 59 [49, 70] | 0.020 |
| Heart rate | 87 [75, 100] | 84 [71, 96] | < 0.001 | 89 [78, 102] | 87 [75, 98] | 0.050 | 90 [79, 103] | 86 [74, 102] | 0.607 |
| SaO2 | 99 [97, 100] | 98 [96, 100] | 0.081 | 99 [97, 100] | 98 [96, 100] | 0.655 | 99 [97, 100] | 99 [97, 100] | 0.104 |
| Respiration rate | 18 [14, 22] | 18 [14, 22] | 0.787 | 17 [14, 21] | 19 [15, 22] | < 0.001 | 18 [14, 22] | 17 [14, 21] | 0.074 |
| Systolic blood pressure | 119 [103, 138] | 116 [102, 132] | 0.008 | 117 [104, 134] | 112 [100, 129] | 0.042 | 118 [104, 134] | 117 [102, 135] | 0.712 |
| Diastolic blood pressure | 61 [54, 70] | 56 [49, 65] | 0.004 | 62 [55, 71] | 57 [51, 65] | < 0.001 | 62 [55, 71] | 60 [53, 69] | 0.806 |
| Mean blood pressure | 80 [70, 92] | 76 [68, 86] | 0.067 | 80 [72, 91] | 76 [67, 86] | 0.017 | 80 [72, 91] | 78 [70, 90] | 0.775 |

**Table V.** Cohort characteristics table for each comparative test in the third matched cohort (MIMIC-III). Continuous variables are represented as medians and IQRs.

|  | African American and Caucasian | Hispanic and Caucasian | Hispanic and African American |
| --- | --- | --- | --- |

| Variables | African American | Caucasian | p-value | Hispanic | Caucasian | p-value | Hispanic | African American | p-value |
| --- | --- | --- | --- | --- | --- | --- | --- | --- | --- |
| Patients (number) | 1333 | 1333 | - | 485 | 485 | - | 484 | 484 | - |
| Gender (male) | 606 (45.46%) | 612 (45.91%) | - | 301 (62.06%) | 301 (62.06%) | - | 303 (62.6%) | 288 (59.5%) | - |
| Age | 61 [51, 72] | 61 [51, 72] | 0.379 | 55 [43, 68] | 55 [43, 68] | 0.765 | 55 [44, 68] | 55 [44, 68] | 0.205 |
| Heart rate | 87 [75, 100] | 85 [74, 97] | 0.810 | 89 [78, 103] | 87 [76, 98] | 0.101 | 89 [78, 102] | 87 [75, 100] | 0.136 |
| SaO2 | 99 [97, 100] | 99 [96, 100] | 0.572 | 99 [97, 100] | 99 [97, 100] | 0.125 | 99 [97, 100] | 99 [97, 100] | 0.576 |
| Respiration rate | 18 [14, 22] | 18 [14, 21] | 0.161 | 17 [14, 21] | 18 [14, 22] | 0.078 | 17 [14, 21] | 18 [14, 22] | 0.066 |
| Systolic blood pressure | 119 [103, 138] | 114 [101, 130] | 0.010 | 117 [104, 134] | 113 [100, 129] | 0.394 | 117 [104, 134] | 121 [105, 140] | 0.446 |
| Diastolic blood pressure | 61 [54, 70] | 57 [50, 65] | 0.287 | 62 [54, 71] | 58 [51, 67] | 0.005 | 62 [54, 70] | 63 [55, 72] | 0.276 |
| Mean blood pressure | 80 [70, 92] | 76 [68, 86] | 0.012 | 80 [71, 91] | 76 [68, 87] | 0.017 | 80 [71, 91] | 81 [72, 93] | 0.176 |

## 2. Appendix 2 Selected variables, measurement units and valid clinical ranges

**Table I.** Variables used in the research, their unit, and the ranges considered during the selection criteria (eICU-CRD).

|  | Patients' general information |  | Patients' medical condition |  | Patient's vital signs |  |  |  |  |  |
| --- | --- | --- | --- | --- | --- | --- | --- | --- | --- | --- |
| Variable | age | gender | diagnosis | patient's status | heart rate | oxygen saturation | respiration rate | systolic blood pressure | diastolic blood pressure | mean blood pressure |
| Unit | years | binary | text | binary | bpm | % | insp/min | mmHg | mmHg | mmHg |
| Range | 18-89 | male/female | / | alive | 0-300 | 0-100 | 0-200 | 0-300 | 0-300 | 0-300 |

## 3. Appendix 3 Full overview of results

**Table I.** AUC and AP across all comparative tests for each matched cohort and each of the three algorithms used (eICU-CRD and MIMIC-III)

|  | African American and Caucasian |  | Hispanic and Caucasian |  | Hispanic and African American |  |  |
| --- | --- | --- | --- | --- | --- | --- | --- |
| eICU-CRD |  |  |  |  |  |  |  |
| Metric | AUC | AP | AUC | AP | AUC | AP | Model |
| Match 1 | 0.62 ± 0.02 | 0.62 ± 0.06 | 0.6 ± 0.05 | 0.57 ± 0.037 | 0.66 ± 0.061 | 0.63 ± 0.066 | LR |
|  | 0.7 ± 0.02 | 0.65 ± 0.069 | 0.62 ± 0.03 | 0.60 ± 0.05 | 0.67 ± 0.038 | 0.63 ± 0.067 | XGBoost Default |
|  | 0.75 ± 0.023 | 0.70 ± 0.092 | 0.66 ± 0.038 | 0.62 ± 0.057 | 0.69 ± 0.043 | 0.64 ± 0.069 | XGBoost Optimised |
| Match 2 | $0.64 \pm 0.016$ | $0.62 \pm 0.059$ | $0.63 \pm 0.056$ | $0.58 \pm 0.042$ | $0.70 \pm 0.051$ | $0.62 \pm 0.064$ | LR |
| | $0.71 \pm 0.023$ | $0.66 \pm 0.071$ | $0.64 \pm 0.04$ | $0.60 \pm 0.062$ | $0.69 \pm 0.041$ | $0.61 \pm 0.053$ | XGBoost Default |
| | $0.75 \pm 0.019$ | $0.70 \pm 0.089$ | $0.67 \pm 0.047$ | $0.63 \pm 0.065$ | $0.72 \pm 0.045$ | $0.65 \pm 0.081$ | XGBoost Optimised |
| Match 3 | $0.62 \pm 0.025$ | $0.62 \pm 0.052$ | $0.61 \pm 0.053$ | $0.58 \pm 0.045$ | $0.66 \pm 0.048$ | $0.61 \pm 0.059$ | LR |
| | $0.71 \pm 0.018$ | $0.66 \pm 0.071$ | $0.61 \pm 0.038$ | $0.58 \pm 0.044$ | $0.67 \pm 0.025$ | $0.60 \pm 0.053$ | XGBoost Default |
| | $0.75 \pm 0.022$ | $0.69 \pm 0.085$ | $0.65 \pm 0.056$ | $0.62 \pm 0.062$ | $0.71 \pm 0.044$ | $0.64 \pm 0.076$ | XGBoost Optimised |
| **MIMIC-III** |  |  |  |  |  |  |  |
| **Metric** | AUC | AP | AUC | AP | AUC | AP | **Model** |
| Match 1 | $0.58 \pm 0.04$ | $0.61 \pm 0.053$ | $0.59 \pm 0.042$ | $0.56 \pm 0.056$ | $0.63 \pm 0.039$ | $0.55 \pm 0.049$ | LR |
| | $0.62 \pm 0.018$ | $0.61 \pm 0.064$ | $0.55 \pm 0.044$ | $0.59 \pm 0.067$ | $0.56 \pm 0.051$ | $0.52 \pm 0.041$ | XGBoost Default |
| | $0.64 \pm 0.024$ | $0.63 \pm 0.071$ | $0.54 \pm 0.038$ | $0.58 \pm 0.052$ | $0.61 \pm 0.035$ | $0.54 \pm 0.062$ | XGBoost Optimised |
| Match 2 | $0.62 \pm 0.02$ | $0.61 \pm 0.055$ | $0.62 \pm 0.033$ | $0.57 \pm 0.048$ | $0.62 \pm 0.036$ | $0.59 \pm 0.06$ | LR |
| | $0.62 \pm 0.017$ | $0.61 \pm 0.067$ | $0.53 \pm 0.035$ | $0.55 \pm 0.040$ | $0.61 \pm 0.035$ | $0.56 \pm 0.05$ | XGBoost Default |
| | $0.65 \pm 0.021$ | $0.64 \pm 0.075$ | $0.60 \pm 0.026$ | $0.58 \pm 0.062$ | $0.64 \pm 0.038$ | $0.58 \pm 0.056$ | XGBoost Optimised |
| Match 3 | $0.62 \pm 0.024$ | $0.60 \pm 0.048$ | $0.62 \pm 0.033$ | $0.57 \pm 0.048$ | $0.63 \pm 0.058$ | $0.59 \pm 0.07$ | LR |
| | $0.61 \pm 0.029$ | $0.61 \pm 0.065$ | $0.53 \pm 0.035$ | $0.55 \pm 0.04$ | $0.59 \pm 0.045$ | $0.56 \pm 0.049$ | XGBoost Default |
| | $0.63 \pm 0.032$ | $0.63 \pm 0.074$ | $0.60 \pm 0.026$ | $0.58 \pm 0.062$ | $0.60 \pm 0.036$ | $0.56 \pm 0.046$ | XGBoost Optimised |

## 4. Appendix 4 AUC-ROC and AU-PRC for the first and third matched cohort across all comparative tests

**Table I.**
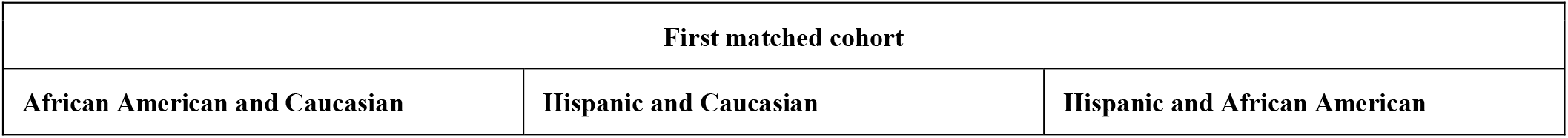
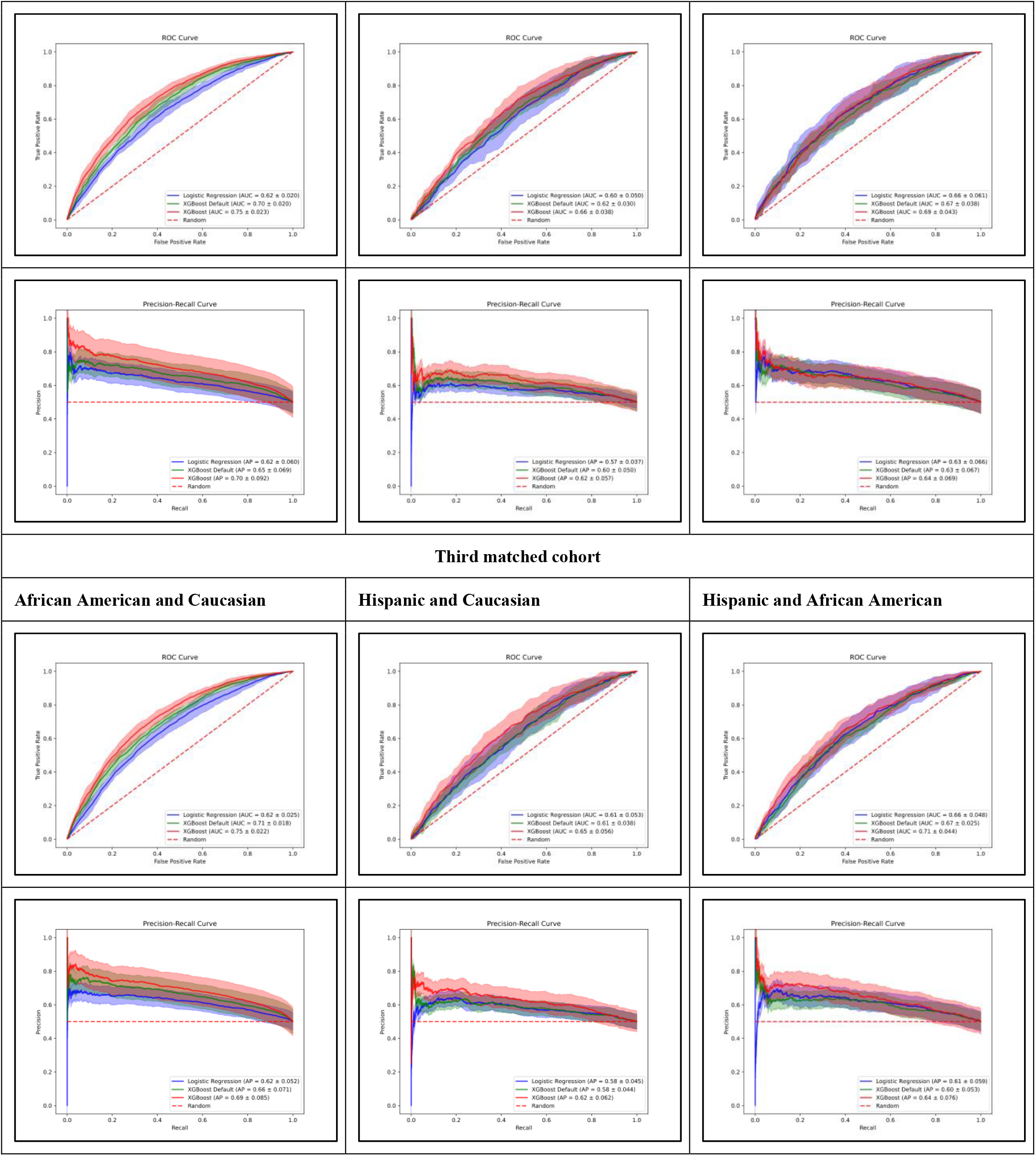
AUC-ROC and AU-PRC for the first and third matched cohort across all comparative tests (eICU-CRD)

**Table II.**
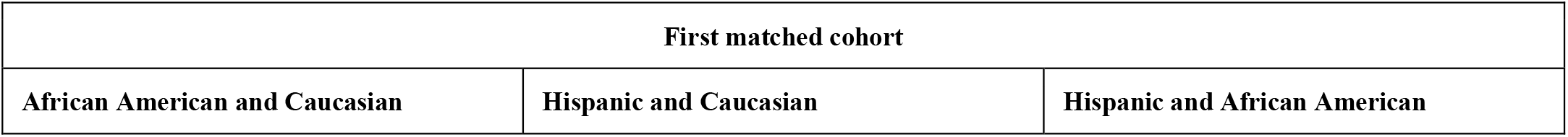
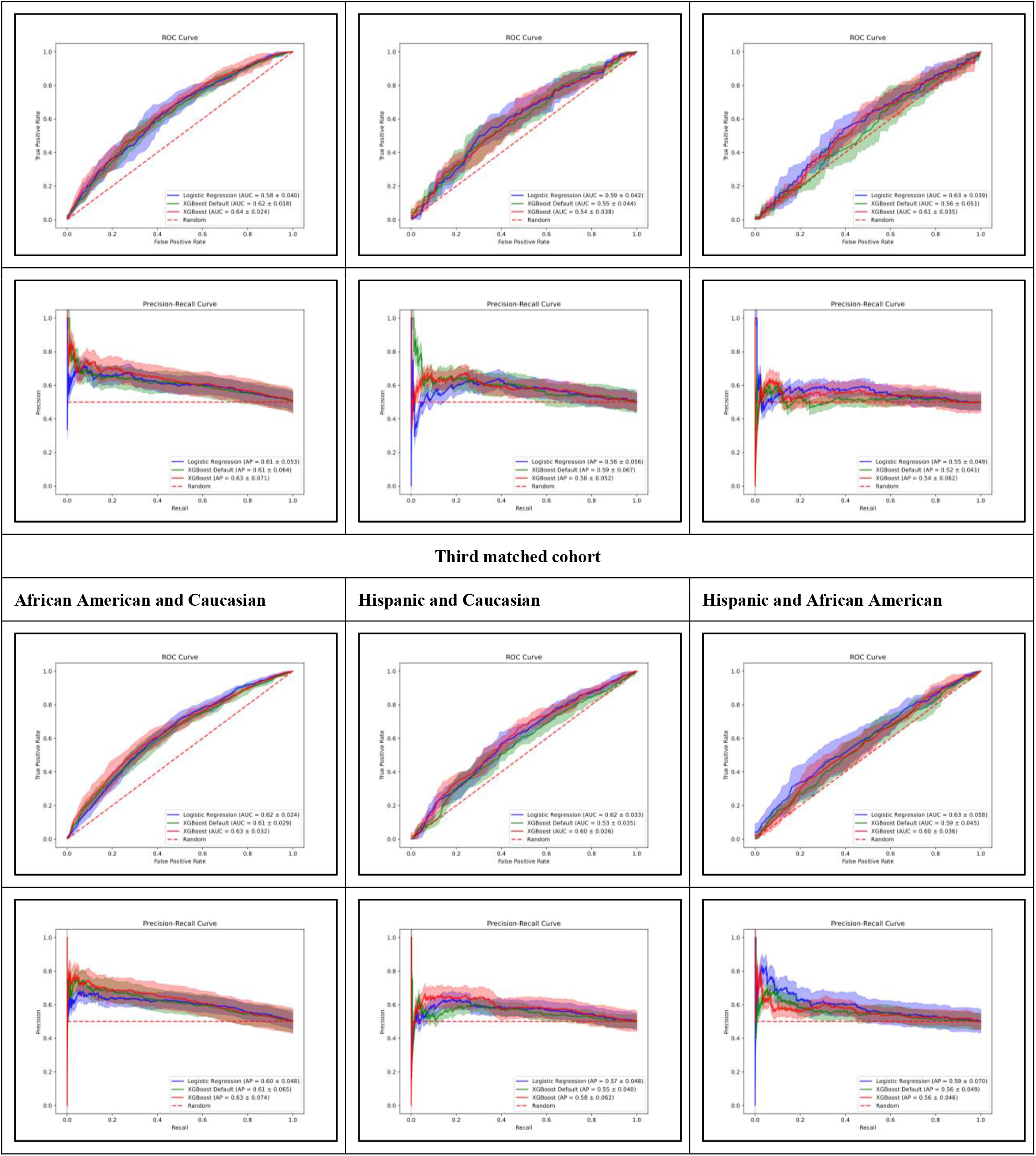
AUC-ROC and AU-PRC for the first and third matched cohort across all comparative tests (MIMIC-III)

